# Evaluation of specimen types and saliva stabilization solutions for SARS-CoV-2 testing

**DOI:** 10.1101/2020.06.16.20133041

**Authors:** Sara B Griesemer, Greta Van Slyke, Dylan Ehrbar, Klemen Strle, Tugba Yildirim, Dominick A Centurioni, Anne C Walsh, Andrew K. Chang, Michael J Waxman, Kirsten St. George

## Abstract

Identifying SARS-CoV-2 infections through aggressive diagnostic testing remains critical in tracking and curbing the spread of the COVID-19 pandemic. Collection of nasopharyngeal swabs (NPS), the preferred sample type for SARS-CoV-2 detection, has become difficult due to the dramatic increase in testing and consequential supply strain. Therefore, alternative specimen types have been investigated, that provide similar detection sensitivity with reduced health care exposure and potential for self-collection. In this study, the detection sensitivity of SARS-CoV-2 in nasal swabs (NS) and saliva was compared to that of NPS, using matched specimens from two outpatient cohorts in New York State (total n = 463). The first cohort showed only a 5.4% positivity but the second cohort (n=227) had a positivity rate of 41%, with sensitivity in NPS, NS and saliva of 97.9%, 87.1%, and 87.1%, respectively. Whether the reduced sensitivity of NS or saliva is acceptable must be assessed in the settings where they are used. However, we sought to improve on it by validating a method to mix the two sample types, as the combination of nasal swab and saliva resulted in 94.6% SARS-CoV-2 detection sensitivity. Spiking experiments showed that combining them did not adversely affect the detection sensitivity in either. Virus stability in saliva was also investigated, with and without the addition of commercially available stabilizing solutions. The virus was stable in saliva at both 4°C and room temperature for up to 7 days. The addition of stabilizing solutions did not enhance stability and in some situations reduced detectable virus levels.

## Introduction

Identifying acute COVID-19 infection through diagnostic testing and screening remains critical in our efforts to care for affected patients and curb the pandemic. Accordingly, laboratory services have worked to expand testing capacity, improve turnaround times, track the course of the outbreak, and provide data for patient management and the implementation of appropriate mitigation efforts and healthcare services (1).

The original standard for diagnosing COVID-19 was molecular testing for SARS-CoV-2 RNA on a nasopharyngeal swab (NPS) collected in virus transport medium (VTM). Unfortunately, performing NPS necessitates the use of a specifically designed swab and requires health care workers to wear extensive personal protective equipment (PPE). Within weeks of the pandemic onset in the US, multiple aspects of the specimen collection and testing process became extremely difficult. The desired testing capacity quickly exceeded anything attempted in the country before and vendors were unable to meet the supply demands for all required sampling components including the specific swabs used for NPS specimen collection, VTM, PPE, reagents, and instruments needed for testing the samples at the laboratories (1-6). In response, a number of changes were implemented: guidelines were rapidly changed to require clinicians to limit the use of (or to reuse) PPE (4, 7-9), alternatives to VTM were tested and implemented (10-12), and numerous additional commercial diagnostic tests and instruments were authorized and distributed by the U.S Food and Drug Administration (https://www.fda.gov/emergency-preparedness-and-response/mcm-legal-regulatory-and-policy-framework/emergency-use-authorization#covid19euas). Additionally, other sample types were assessed as alternatives to NPS. (3, 13-19)

Testing of oropharyngeal swabs (OPS), nasal swabs (NS), mid-turbinate swabs (MTS) and saliva provided the opportunity to be less restrictive about the type of swab used, and the last three options also offer the additional advantage of self-collection. This has been demonstrated to be successful in previous studies on influenza detection, where self-collected specimens were found to have similar detection rates as those collected by health care personnel (20, 21). Major advantages of self-collection include limiting the exposure of health care workers to potentially infected patients, subsequently reducing usage of PPE, as well as limiting the exposure of patients to hospitals where infection is likely more prevalent. Self-collection also allows for individuals to conveniently test themselves at home, which could lead to increased testing rates with a limited burden on health care systems.

Numerous recent studies have therefore focused on the comparative sensitivity of upper respiratory swabs other than NPS, which has long been considered the preferred option for optimal detection of most respiratory viruses (3, 15, 19, 22). Experience with saliva as a diagnostic sample type for respiratory viruses has been limited, although in recent weeks, several studies have suggested saliva to be of acceptable or equal sensitivity to NPS or other specimen types for the diagnosis of SARS-CoV-2 (14, 17, 18, 23, 24). Some studies have also proposed the use of saliva stabilizing solutions and it has been unclear as to whether these are necessary or advantageous.

In this study, nasal swabs and saliva were compared to NPS for the detection of SARS-CoV-2 in outpatients from two distinct New York populations, one of which had high prevalence while the other had low. The stability of SARS-CoV-2 in saliva and the utility of three stabilizing solutions were investigated. Finally, methods for combining swabs and saliva in the laboratory were assessed to maximize detection of the virus without running multiple tests when more than one specimen type is collected.

## METHODS

### Specimen collection sites and patients

This study was granted non-research determination under the emergency response criterion by the New York State Department of Health Institutional Review Board.

Two sites were accessed for specimens collected between March 20 and March 26, 2020. The first testing site was a large tent erected on the Albany Medical Center (AMC) Emergency Department parking lot in Albany, NY. Open from 7am to 11pm for 2.5 days, this walk-in site enrolled 236 subjects. Patients without an appointment could be tested for COVID-19 and were not required to be symptomatic. No patients were excluded. The second testing site was a drive-through Regional Operations Center in New Rochelle (NR), NY, where an identified “super-spreader” event had taken place early in the pandemic and the COVID-19 positivity rate was known to be high. Patients were required to have an appointment and qualifying symptoms or exposures; 227 subjects were enrolled from this site.

### Specimen types, collection and transport

Three specimens were collected from each subject: nasopharyngeal swab, nasal swab, and saliva. Nasal swab and NPS were placed in separate tubes containing 1 ml Molecular Transport Media (Longhorn Vaccines and Diagnostics, LLC., San Antonio TX). Saliva samples were collected in sterile 50mL conical tubes, and patients were instructed to refrain from eating, drinking, chewing gum or tobacco, or smoking, 30 minutes prior to collection. Specimens from AMC were brought to the Wadsworth Center Laboratory of Viral Diseases (LVD) approximately every 2-4 hours for accessioning and testing. Those from NR were placed in coolers and transported to the LVD within 24 hours for accessioning and testing. Specimens were held at 4°C from the time of collection to the time of processing into lysis buffer for molecular testing. All testing was performed within 24-72 hours of the time of specimen collection.

### SARS-CoV-2 Testing

Saliva specimens with excessive mucus were digested prior to extraction with Snap n’ Digest (Scientific Device Laboratory, Des Plaines, IL); briefly, 0.5-1 ml of Snap n’ Digest was added to each saliva specimen based on sample volume, vortexed and allowed to digest for 10 minutes. The CDC 2019 nCoV Real-Time RT-PCR Diagnostic Panel (25-27) was used to test all samples. Total nucleic acid extraction was conducted on the bioMerieux easyMAG^®^ or EMAG^®^ (bioMerieux Inc, Durham, NC); briefly, for all individual specimens tested, 110μL of the sample was added to 2mL NucliSENS Lysis Buffer (bioMerieux) and extracted into 110μL of eluate. Testing was performed on ABI 7500FAST Dx instruments as described in the instructions for use (IFU) on the FDA website. All cycle threshold (Ct) values for viral RNA detection presented herein are for the N1 gene.

### Virus and viral RNA quantification

SARS-CoV-2 isolate, USA-WA1/2020, was obtained from BEI Resources (NIH/ATCC Manassas, VA) and amplified in Vero E6 cells, in accordance with Biosafety Level 3 procedures (BSL-3). Viral isolate RNA was extracted and quantified by standard curve using RNA transcript synthesized to contain all SARS-CoV-2 N-gene targets in the CDC 2019-nCoV Real-Time RT-PCR Diagnostic Panel (bio-synthesis, Lewisville, TX).

### Validation of different specimen types for viral detection

Validation of all specimen types with the CDC 2019-nCoV Real-Time RT-PCR Diagnostic Panel included a limit of detection determination, followed by a clinical evaluation panel. In order to perform these experiments in a BSL-2 environment, quantified SARS-CoV-2 viral RNA, rather than infectious virus, was spiked into lysed negative specimens prior to extraction. Saliva was validated for use on the easyMAG and EMAG instruments, as well as the MagNA Pure 96 system (Roche Diagnostics, Indianapolis, IN). Combined saliva and nasal swab specimens were validated on the easyMAG and EMAG instruments only. On the MagNA Pure 96, 100uL specimen was added to 350uL lysis buffer and eluted into 100uL.

### Limits of detection

Limits of detection (LOD) were determined by testing a range finding dilution series in triplicate to estimate the LOD, then confirmed by spiking that concentration of SARS-CoV-2 viral RNA into 20 individual clinical specimens. The LOD was assessed as that which gave at least 95% detection of the sample replicates.

### Combined saliva and nasal swab as a single specimen

To investigate the possibility of maximizing virus detection without doubling the number of tests performed, mixed saliva and nasal swabs were validated as a sample type in the following manner: 110uL of each specimen was added to the same 2mL lysis buffer tube, eluted into 110uL and tested as one sample. To mimic situations in which saliva is positive and nasal swabs are negative, or vice-versa, and confirm that there is no adverse impact on the detection, LODs were determined for spiked combined specimens.

### Clinical evaluations

To evaluate the detection of SARS-CoV-2 in clinical saliva samples, individual negative saliva specimens were spiked with quantified viral RNA at the following range of concentrations in triplicate: 2X, 4X, 6X, 8X, 10X, and 100X the established LOD. These evaluations were also performed with combined saliva and nasal swab specimens. Additionally, 19 previously positive nasal swabs in Viral Transport Media (VTM), with original Ct values ranging from 20-39, and three positive saliva specimens, with Ct values of 21-33, were individually re-analyzed and tested in combination with the other negative samples.

### Saliva stabilizing experiments

To evaluate the stability of SARS-CoV-2 in saliva and to determine the necessity of stabilizing solutions for virus preservation in saliva, infectious SARS-CoV-2 virus was spiked into pooled negative saliva and sampled at multiple timepoints, with storage at multiple temperatures, with and without the addition of various stabilizing buffers. Individual saliva specimens were tested for SARS-CoV-2 by real-time RT-PCR before using in stability studies to verify their negative status. Table 1 summarizes the multiple parameters assessed in the stabilizing experiments. SARS-CoV-2 isolate USA-WA1/2020, cultured and harvested as described above, was used in all stabilizing experiments. Stock viral RNA concentration was calculated at 2.2×10^7^ RNA gene copies/uL as described above. Stock virus was diluted in EMEM + 2% FBS to working concentrations and samples in each experiment were spiked with either high (1×10^6^ RNA copies/mL specimen or 5000 RNA copies/PCR reaction) or low (2×10^5^ RNA copies/mL specimen, or 200 RNA copies/PCR reaction) concentrations in duplicate. Individual saliva specimens were tested for SARS-CoV-2 by real-time RT-PCR before using in stability studies. Sampling of all specimens for the stability studies included lysis of 110uL specimen into 2mL NucliSENS lysis buffer, extraction on EMAG or easyMAG instruments into 110uL of elution buffer, and analysis by the CDC Real-Time RT-PCR Diagnostic Panel. Ct values were averaged and plotted using GraphPad Prism software version 8.3.0.

**Table 1:** Experimental parameters for stability studies

| Matrix | Virus Concentration Spiked (gene copies) | Temperature | Timepoints |
| --- | --- | --- | --- |
| Saliva only<br>VTM only<br>Nasal Swab only<br>Saliva + Nasal Swab only<br>Saliva + VTM<br>Saliva + 23andMe® stabilization buffer<br>Saliva + AncestryDNA® stabilization buffer<br>Saliva + AncestryDNA® stabilization buffer, combined with nasal swab | High: $1 \times 10^6$ gc/mL<br>Low: $2 \times 10^5$ gc/mL | 4°C<br>Room Temp<br>-80°C | Day 0<br>Day 1<br>Day 3<br>Day 5<br>Day 7 |

### 23andMe^®^ stabilization buffer

Saliva stabilization kits were kindly donated by 23andMe^®^ (Sunnyvale, CA). Stabilizing buffer was removed from the kits and pooled. A total of 32 tubes (15ml polypropylene), each containing 1mL saliva, were spiked with SARS-CoV-2 (16 high and 16 low virus concentrations), followed by the immediate addition of 1mL stabilization buffer, then mixed thoroughly. An additional 32 tubes were prepared containing 0.5mL saliva spiked with high and low virus, followed by immediate addition of 0.5mL VTM, then mixed thoroughly. All samples were then sampled and analyzed by real-time RT-PCR (Day 0) by standard procedures, starting with 110uL transfer to lysis buffer tubes as described above. Duplicate tubes at each concentration were then stored at room temperature, 4°C, and −80°C, and samples tested at each of days 1, 3, 5 and 7, except for those stored at −80°C, which were only analyzed at day seven.

### AncestryDNA^®^ stabilization buffer

AncestryDNA^®^ saliva collection kits were kindly donated by the company (Lehi, Utah) and used for additional saliva stability studies. In these experiments, the stability of SARS-CoV-2 was tested in saliva, VTM, saliva with VTM, and saliva with AncestryDNA^®^ (AcD) stabilization buffer. Briefly, tubes containing SARS-CoV-2 negative saliva or VTM were spiked with high and low virus concentrations (six replicates each) followed by the addition of stabilization buffer, mixing, and sampling as above, for Day 0 analysis. Duplicate tubes of each concentration were prepared for each storage temperature and stored at room temperature, 4°C, and −80°C for up to seven days. Stored, spiked samples were tested at appointed days when a 110uL aliquot from each tube was lysed, extracted and tested by real-time PCR at days 1, 3, and 7, except for those stored at −80°C, which were only sampled at day seven.

### Stabilization with saliva and nasal swab combination

AncestryDNA® saliva collection kits were used for stability studies in an experiment to investigate the stability of SARS-CoV-2 in saliva, nasal swab in VTM, and saliva with AcD stabilization buffer. In addition, to observe the effects of combining specimens on SARS-CoV-2 detection sensitivity, saliva alone and in combination with nasal swabs in VTM were lysed and analyzed together. Pooled negative saliva and pooled nasal swabs in VTM, all negative for SARS-CoV-2, were used in this experiment. A total of 18 tubes were prepared, 12 containing saliva (alone) and 6 containing NS in VTM. Four saliva tubes were spiked with high virus, four with low virus and four remained un-spiked. Two of each of these four (2 high, 2 low, 2 no virus) were then mixed with AcD stabilization buffer. Six NS tubes were also spiked with high, low, or no virus. Tubes were incubated at room temperature and sampling was performed at three timepoints: days 0, 1.5, and 3. At each time point, 110uL from each of the tubes was removed, lysed, extracted and analyzed. In addition, 110uL of each prepared saliva sample, with and without stabilization buffer, was lysed together with 110uL of each prepared nasal swab sample, so that high, low, and negative saliva samples would be mixed with high, low, and negative nasal swab samples. Combined samples were then extracted as one specimen and analyzed.

### Statistical analyses

For all statistical calculations, each Ct value used is the mean of the N1 and N2 Ct values for that positive specimen. In cases where a specimen type was not collected for an individual, that patient was then excluded from calculations of that specimen type’s sensitivity. For the purpose of calculating sensitivities, true positives were determined as any Ct value < 45 for any specimen type. Sensitivities and associated confidence intervals were calculated using the R package pROC (28). Confidence intervals of sensitivities were calculated using 2000 stratified bootstrap replicates. Two-sample Wilcoxon rank-sum tests were used to compare mean ranks of Ct values between males and females for each specimen type. Wilcoxon rank-sum tests were performed, and the results plotted, using the R package ggpubr (Alboukadel Kassambara (2020). ggpubr: ‘ggplot2’ Based Publication Ready Plots. R package version 0.3.0). Conditional density plots were created using the R package vcd (David Meyer, Achim Zeileis, and Kurt Hornik (2020). vcd: Visualizing Categorical Data. R package version 1.4-7) with the default settings. R version 3.6.3 (R Core Team (2020). R: A language and environment for statistical computing. R Foundation for Statistical Computing, Vienna, Austria) was used for all preceding analyses.

## RESULTS

### Patient demographics and regional positivity rates

Specimens were collected from a total of 463 individuals at two collection sites in New York State during the early weeks of the SARS-CoV-2 pandemic. Samples were initially collected from 236 individuals at AMC. However, following initial testing of NPS, only 12 of 236 patients tested positive for SARS-CoV-2 RNA, a positivity rate too low (5%) to enable statistically meaningful comparisons (Figure 1A). Specimen collection was immediately transferred to a location with known high positivity. During March 2020, New Rochelle (NR), NY, was one of the Northeast’s earliest pandemic “hot spots” with a significantly higher positivity rate than the rest of the state. Specimens were collected from 227 patients in NR with demographics as shown in Table 2. Of the 227 patients tested at that site, 93 (41%) were positive for SARS-CoV-2, with 74 patients positive in all three sample types, 7 positive in both NS and NPS, 5 positive in both saliva and NPS, and 5 positive in NPS alone. Interestingly, 2 patients were positive in saliva alone but none in NS alone, while NPS detected 91 of 93 positive patients (Figure 1B). When Wilcoxon rank-sum tests were used to compare mean ranks of Ct values between males and females for each specimen type, no significant difference in positivity rates were seen between genders (data not shown). The distribution of overall positivity by age for the NR cohort is shown in Figure 2. Linear regression was used to examine the relationships between Ct values in each specimen type and the age of the patients. Only small, but significant, negative correlations between Ct values from NPS and saliva specimens with age were found (data not shown).

**Figure 1.**
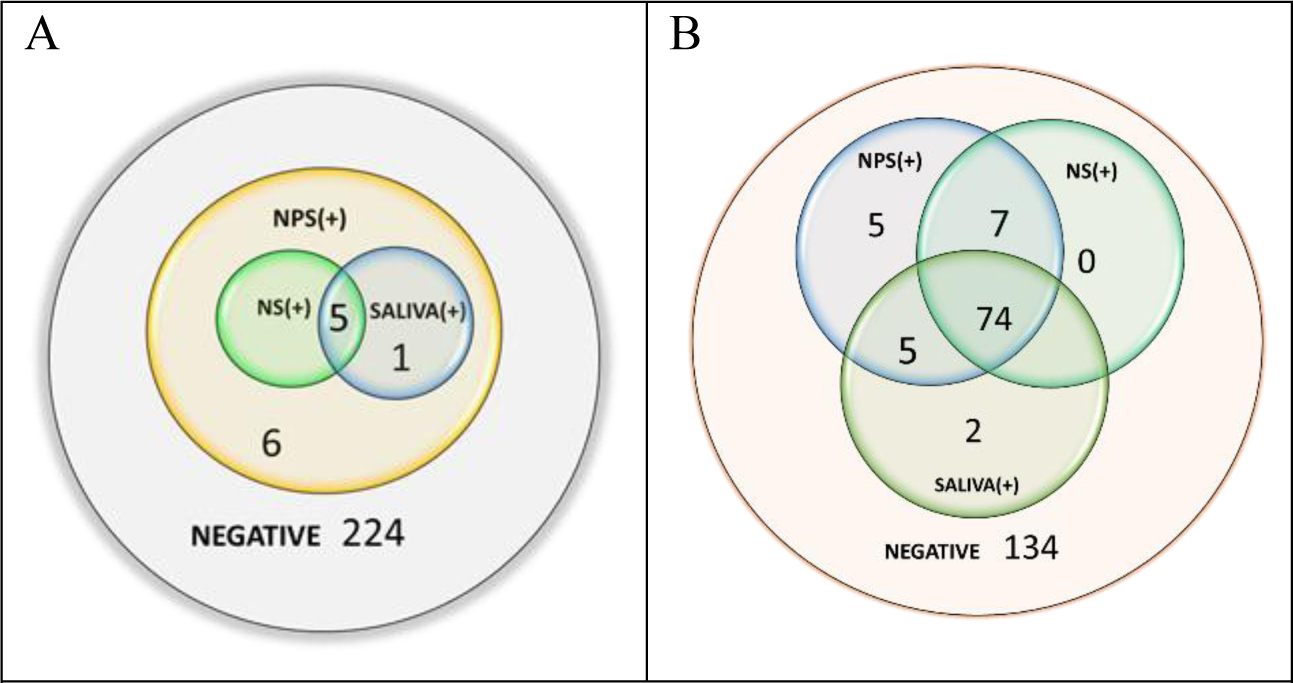
Comparison of positivity in different specimen types from the two collection sites: A. Albany Medical Center (n=236) and B. New Rochelle (n=227).

**Table 2:** Patient Demographics

|  | Albany Medical Center Cohort |  | New Rochelle Cohort |  |
| --- | --- | --- | --- | --- |
|  | All patients (N=236) | Positive patients (N=12) | All patients (N=227) | Positive patients (N=93) |
| Gender, male | 47% | 25% | 60% | 60% |
| Age, years |  |  |  |  |
| 0-5 | 4 | 0 | 0 | 0 |
| 6-18 | 13 | 0 | 2 | 1 |
| 19-45 | 133 | 6 | 149 | 63 |
| 46-65 | 68 | 6 | 59 | 21 |
| >65 | 16 | 0 | 6 | 4 |
| Unknown | 2 | 0 | 11 | 4 |
| Age range | 3-105 | 27-61 | 14-77 | 14-71 |

**Figure 2:**
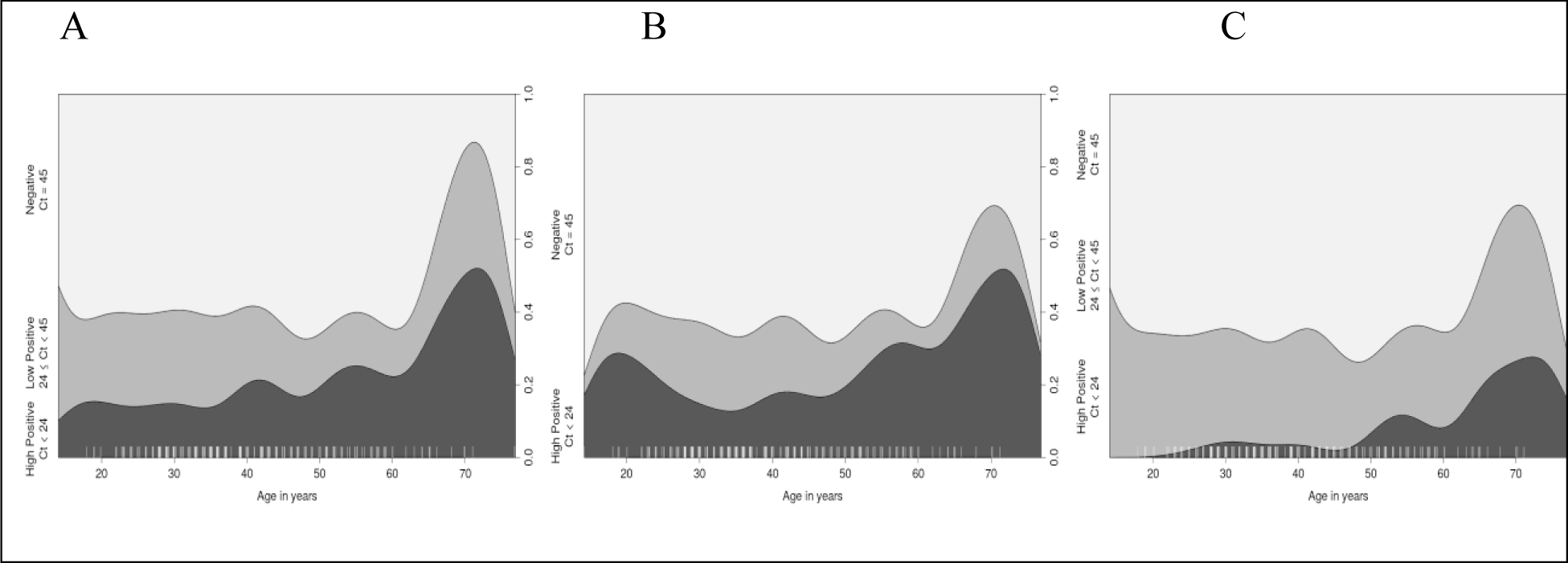
Distribution of negative, low positive, and high positive patients by age. Conditional density plots displaying the distribution of negative, low, and high positive samples by age. High positive samples (darkest grey) showed Ct values < 24, low positives (mid grey) had 24 ≤ Ct < 45, and negatives (light grey) had Ct values of 45. Results are shown for (A) NPS, (B) NS, or (C) saliva.

### Comparison of specimen sensitivities for SARS-CoV-2 detection

We sought to determine whether NS or saliva specimens could be used as a surrogate sample type in place of NPS for SARS-CoV-2 detection when using the CDC 2019 nCoV Real-Time RT-PCR Diagnostic Panel. Comparisons of grouped Ct values using a Kruskal-Wallis test with Dunn’s multiple comparisons post-hoc test revealed statistically significant differences between the three sample types: the mean Ct values for both NPS and NS positive samples was significantly higher (P<0.0001) than that of positive saliva samples and the mean Ct values for NPS and NS were not significantly different from one another (Figure 3). When sensitivities and corresponding 95% confidence intervals for all specimen types were calculated, NPS and the combination of NS and saliva provided the highest sensitivities (97.8% and 94.6%, respectively) with overlapping 95% confidence intervals (Table 3). Both NS and saliva had lower, and identical, sensitivities and 95% confidence intervals (87.1% and 79.57-93.55 for both) than either NPS or the combination of NS and saliva. The confidence intervals for NS and saliva overlapped with that for the NS and saliva combination specimen type, but not with NPS specimen type (Table 3).

**Figure 3.**
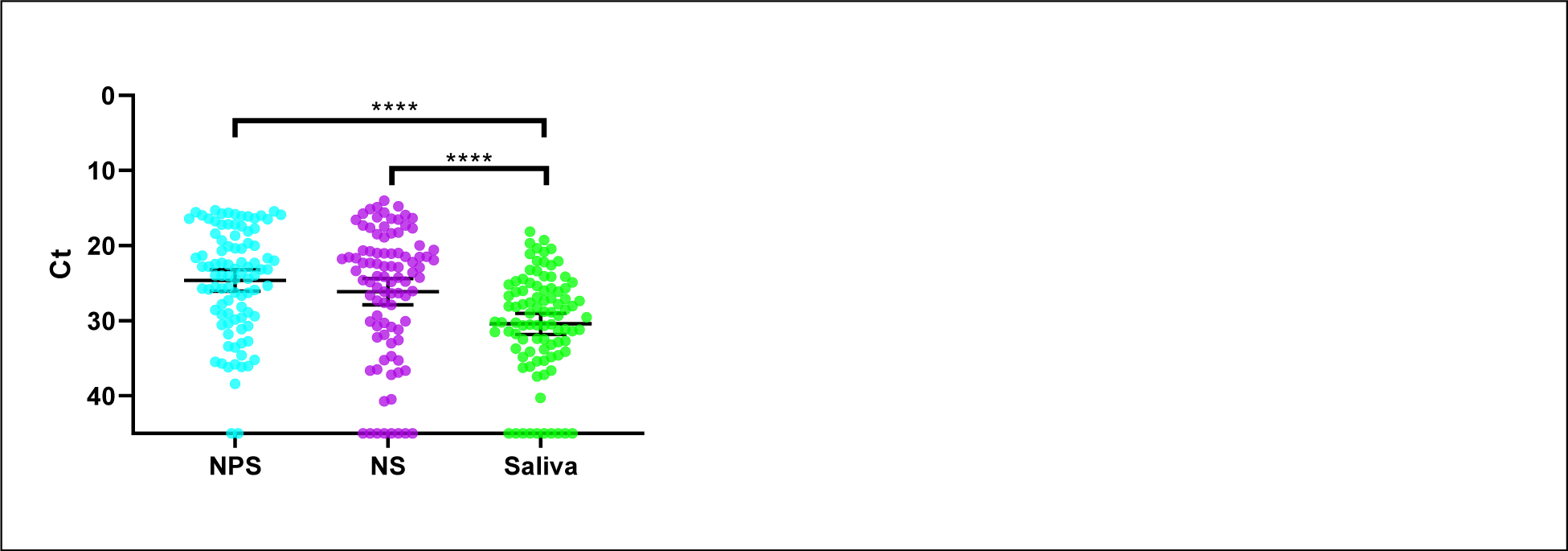
Comparison of SARS-CoV-2 detection by sample type and Ct value. Matched Ct values for NPS, NS, and saliva from all positive individuals collected at the ROC (n= 93) were compared using a Kruskal-Wallis test (P<0.0001) with Dunn’s multiple comparisons post-hoc test. Bars represent mean and 95% CI, and a Ct value of 45 was assigned to undetected specimens

**Table 3:** Sensitivity of detection for NPS, NS, saliva and NS + saliva

| Specimen type | Sensitivity (95% CI) |
| --- | --- |
| NPS | 97.85 (94.62 – 100) |
| NS | 87.10 (79.57 – 93.55) |
| Saliva | 87.10 (79.57 – 93.55) |
| NS and Saliva | 94.62 (89.25 – 98.92) |

### Validation of Saliva and Nasal swabs for clinical testing

Saliva and NS as individual specimen types and the combination of NS + saliva, were all validated for testing on the CDC 2019 nCoV Real-Time RT-PCR Diagnostic Panel after extraction on the EMAG and easyMAG. The LODs for saliva, NS, and NS + saliva, were all found to be 25 RNA copies/PCR reaction or 5 RNA copies/µL on both extraction platforms. Additionally, clinical evaluation studies of all specimen types with amended negative samples showed 100% accuracy (data not shown).

A total of 20 individual negative saliva specimens were separately lysed and spiked with a concentration of 25 RNA copies/PCR reaction. Then, 20 individual negative nasal swabs, in VTM, were added to the lysed saliva and the combined sample analyzed. For the reverse, 20 individual negative nasal swab specimens were separately lysed and spiked with 25 copies/PCR reaction and the same volume of 20 negative saliva samples was added to the lysed nasal swabs. These combined samples were then extracted and analyzed.

A clinical evaluation was also performed on samples of mixed NS and saliva specimens from positive patients. A total of 15 previously positive NS samples were retrieved and retested with and without the addition of saliva. All were positive when combined and Ct values were not adversely affected. (Figure 4). Linear regression analysis indicated a high correlation between nasal swab only Ct values, and nasal swab with saliva Ct values, even for samples with moderately weak values (Ct values between 30-35).

**Figure 4:**
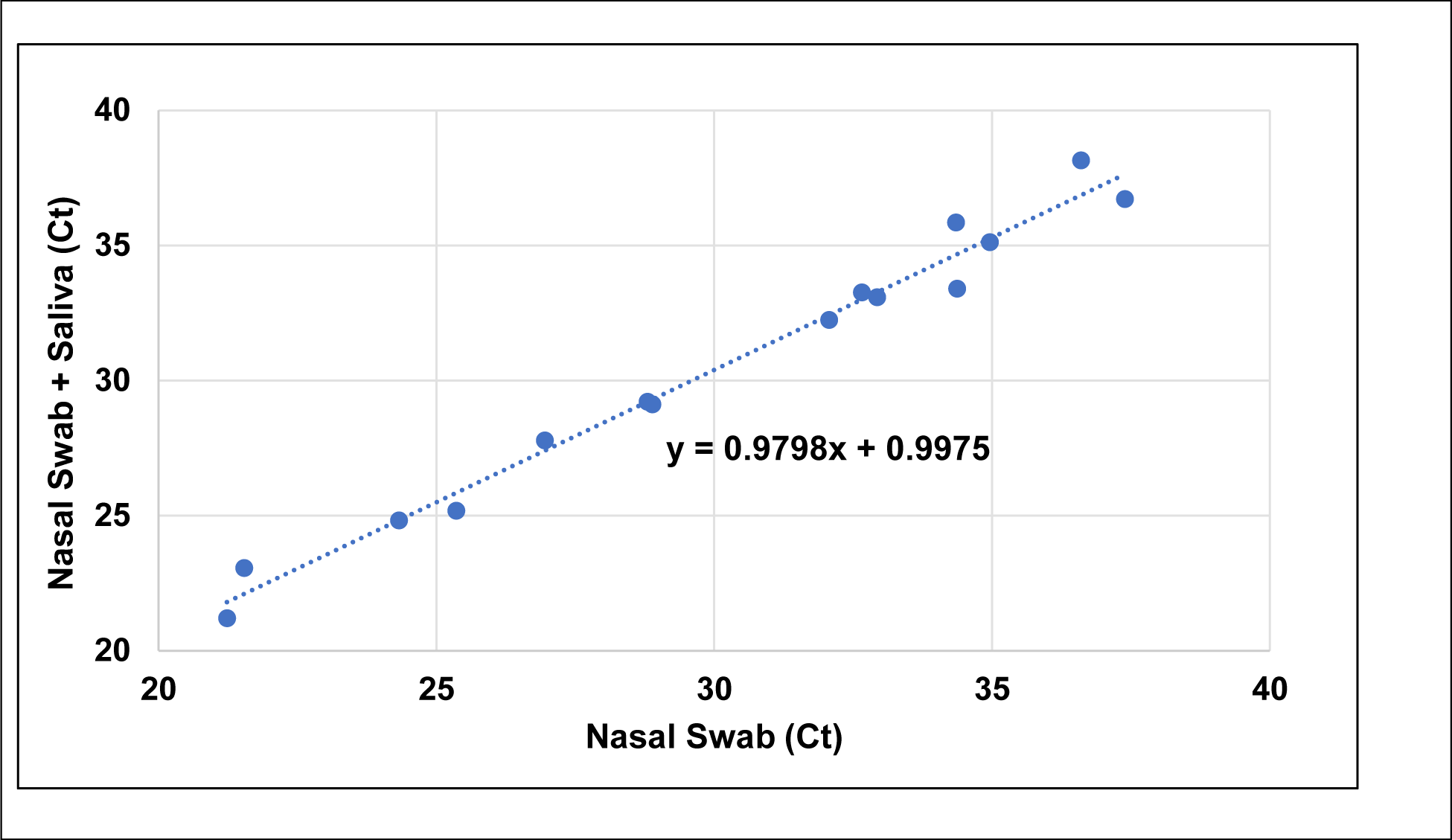
Addition of saliva does not interfere with Ct values of nasal swabs. Nasal swab specimens, previously positive for SARS-CoV-2, were retrieved from −80°C storage and retested by real-time RT-PCR. Negative saliva was extracted together with the same nasal swab specimens. Linear regression analysis was performed on both comparisons.

### SARS-CoV-2 Stability in Saliva with Stabilization Solutions

A series of experiments were then performed in which virus-spiked saliva was supplemented with different stabilization buffers, to examine the stability of SARS-CoV-2 in saliva and the utility of commercial stabilizing solutions. Samples were tested at high and low virus levels, held at different temperatures for various time periods (as described in Materials and Methods), and analyzed for changes in Ct values that would indicate a loss of detectable viral RNA. An initial experiment compared the deterioration of virus in spiked saliva, stabilized by the addition of VTM, to that of spiked saliva with the stabilization buffer from 23andMe^®^. Samples were stored at 4°C and room temperature and tested at days 0, 1, 3, 5 and 7. The SARS-CoV-2 virus in saliva was more stable in VTM than in 23andMe^®^ stabilizing solution, as indicated by Ct values over time at either temperature (Figure 5). Greatest losses in detectable viral RNA were seen at day 7 in saliva with 23andMe^®^ stabilizing solution, particularly in samples that had been stored at room temperature, while those in VTM showed little to no change. A similar experiment comparing viral RNA detection in saliva supplemented with VTM or stabilization solution from Ancestry^®^ also showed no benefit from the addition of stabilization solution and again a significant loss of detectable virus at day 7 in samples that had stabilization solution added to them. Interestingly, in this experiment the virus in spiked saliva with no additives was just as stable as that in VTM, producing unchanged Ct values after 7 days storage at either 4°C or room temperature (Figure 6). The virus was also just as stable in NS at either 4°C or room temperature as in saliva over a three-day test period, while again losing detectable virus in saliva if stabilization buffer was added (Figure 7).

**Figure 5:**
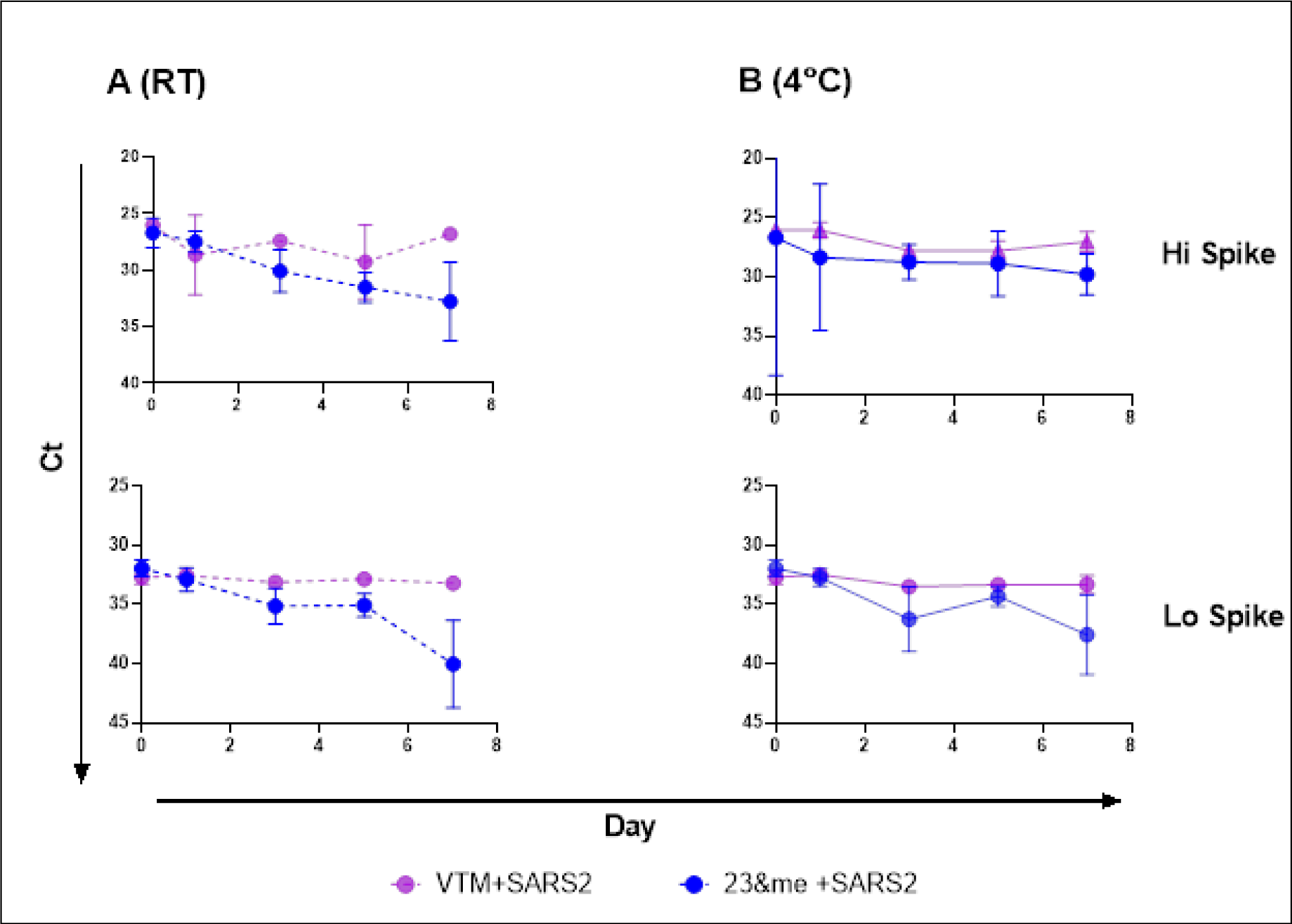
Virus stability in saliva samples mixed with 23andMe^®^ stabilization buffer or VTM. Mixtures of saliva plus 23andMe^®^ buffer (blue) or VTM (purple) and virus were held at either room temperature (A, dashed lines) or 4°C (B, solid lines) and sampled at days 0, 1, 3, 5, and 7, in duplicate. Mean Ct values for N1 were plotted, and simple linear regression analysis performed. The slope of all VTM curves did not deviate significantly from zero; the slope of 23andMe^®^ lo spike at 4°C did not deviate significantly from zero, the slope of all other 23andMe^®^ plots significantly deviated from zero (P<0.02).

**Figure 6:**
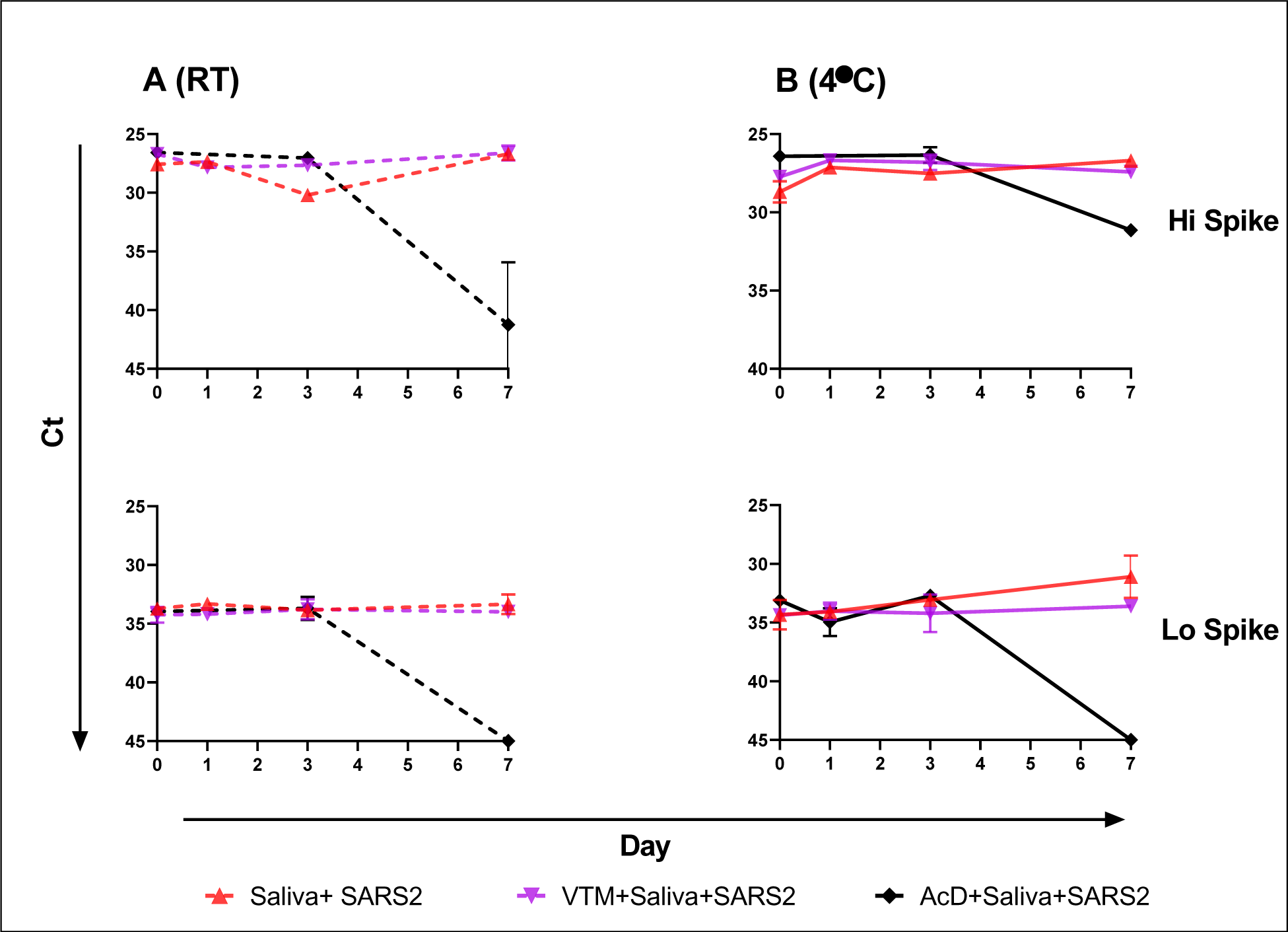
Virus stability in saliva alone, saliva with VTM or saliva combined with AncestryDNA^®^ (AcD) stabilization buffer. Saliva alone (red), saliva plus VTM (purple), and saliva plus AcD buffer, all spiked with virus, were held at room temperature (A, dashed lines) or 4°C (B, solid lines) and sampled, in duplicate, at days 0, 1, 3, and 7. Mean Ct values were plotted, and simple linear regression analysis performed. Neither slopes for saliva alone nor saliva plus VTM deviated from zero; all slopes for saliva plus AcD buffer significantly deviated from zero (P<0.02).

**Figure 7:**
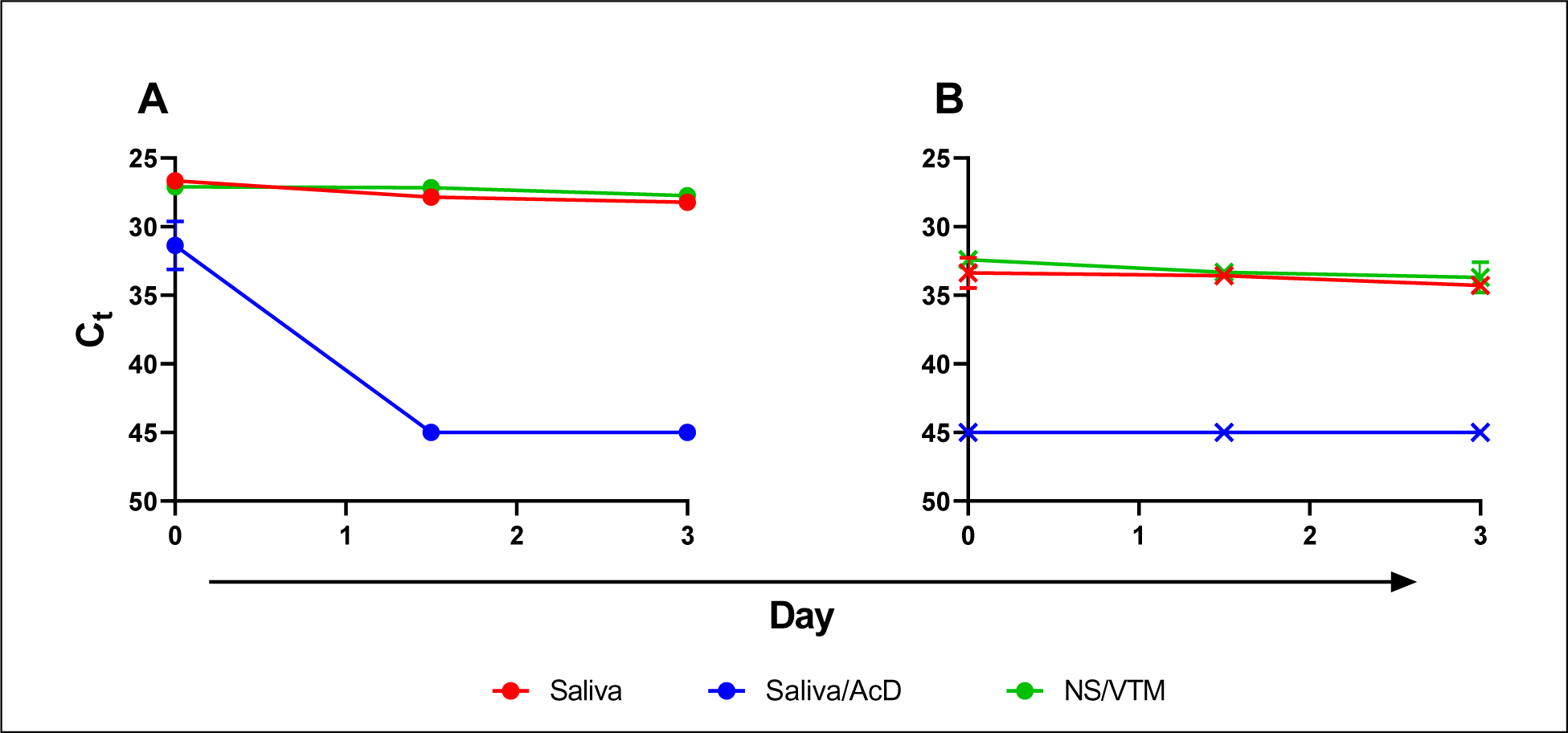
Virus stability over time. Viral RNA detection at both high (A) and low (B) spiked virus concentrations in saliva (red), saliva plus AcD buffer (blue) and, NS in VTM (green). Samples were held at RT and collected, in duplicate, at 0, 36, and 72 hours; mean Ct values are plotted.

In a final series of experiments, we explored whether adding saliva and nasal swabs together and testing as a single specimen impacted the detection of the virus, and whether stabilization solution being previously added to the saliva impacted detection when stored at room temperature for up to three days. Results are graphed in Figure 8 and show that A) the presence of a high virus level in saliva is not impacted by the addition of a high, low, or negatively spiked nasal swab, B) a low level spiked saliva is not impacted by a low or negatively spike nasal swab and the addition of a high level spiked nasal swab raises the virus level to high (detecting the high level virus in the nasal swab), C) the presence of saliva does not impact the detection of low or high level virus spiked into nasal swab. All these findings are consistent across the three days that the sampling was performed. Interestingly, when the same sample combination experiments are performed using saliva supplemented with stabilization buffer, the presence of the solution does not affect the detection of the virus in nasal swabs, however, it does affect its detection in saliva (Figures 8 D, E, and F). With the exception of the detection of some high level spiked virus in saliva at time point 0 (Figure 8, D) the addition of stabilization buffer effectively eliminated the virus detection in saliva at all other time points and concentrations (Figure 8, D and E) leaving detectable virus only in the NS.

**Figure 8:**
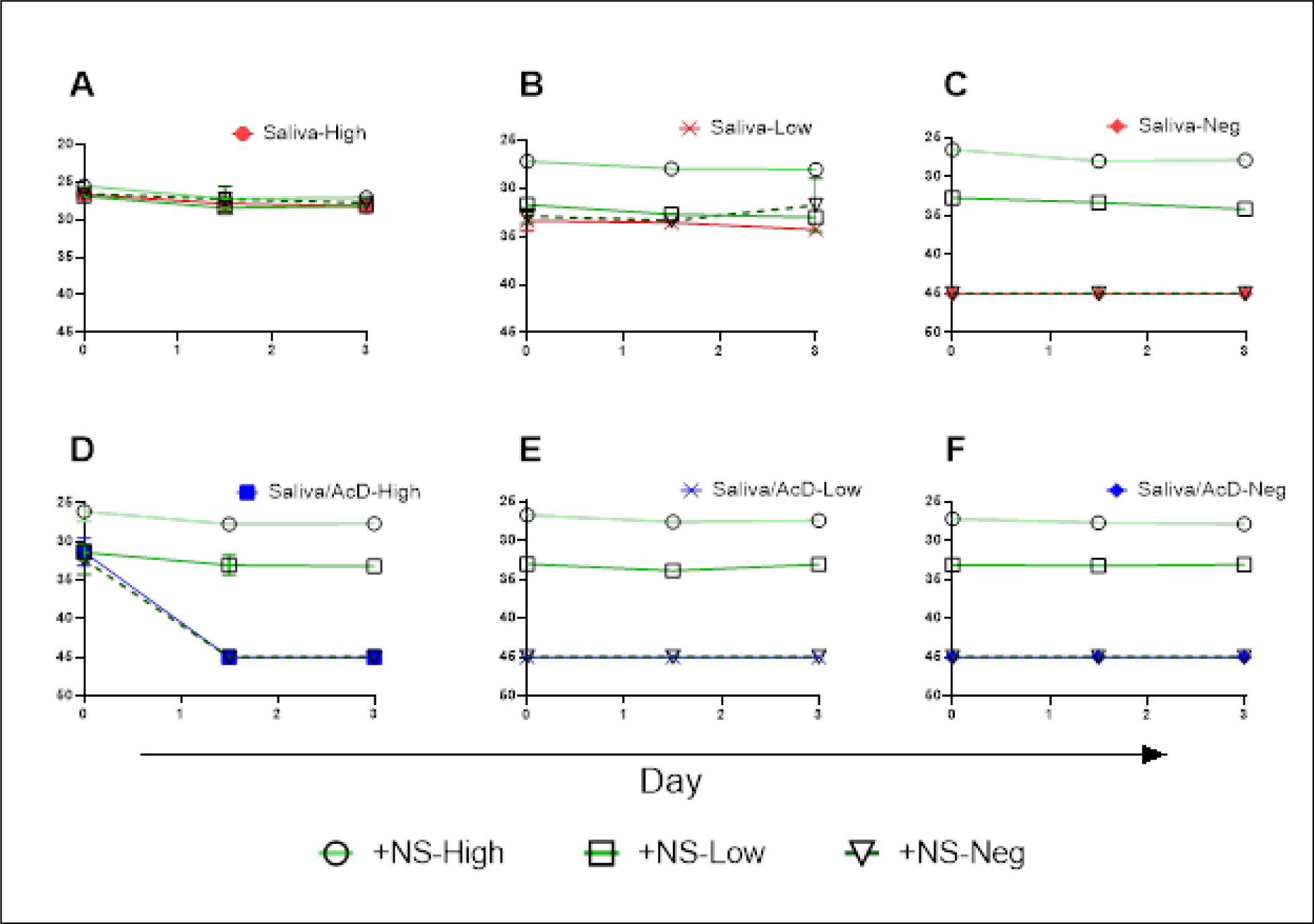
The effect of mixed saliva and NS specimens with high and low virus concentrations, with and without the addition of stabilization buffer to the saliva. Figures A, B, and C show high, low and neg saliva, mixed with high, low and neg NS. Figures D, E, and F show high, low and neg saliva with stabilization buffer, mixed with high, low and neg NS.

## DISCUSSION

As the COVID-19 pandemic continues, ongoing supply issues with collection devices and PPE at a national and international level, together with increasing demands for laboratory testing, has necessitated the exploration of alternative options for specimen types in order to maintain effective mitigation strategies. It will be difficult to find a consensus for the relative performance of different sample types across studies performed on different patient groups for a disease with such variable severity and presentations, across a wide range of age groups, using an array of collection devices and transport media, and with the additional variables of time post-onset and time post-sample collection.

In the study reported here, the results show that in the outpatient group tested, that is not severely ill, NPS specimens produced greatest sensitivity at approximately 98%, while NS or saliva alone provided sensitivities of approximately 87%. Whether the reduction in detection sensitivity that occurs with the use of these specimen types is acceptable has to be assessed in the settings in which they are being used, taking into account the more ready availability of NS swabs, the convenience of collection, the reduced discomfort of NS and saliva collection, as well as the potential for self-collection of these specimens and thus diminished risk of transmission to health care personnel. Notably, some other studies have not found sensitivity differences this great, while others have found them more extreme (13, 14, 16, 18, 19, 24, A. L. Wylie et al. submitted for publication). By combining the NS and saliva specimens in the laboratory, we were able to increase sensitivity to 95%, an additional 8% above NS or saliva alone; we believe this to be a substantial improvement and a beneficial option for sensitive diagnosis of SARS-CoV-2.

Of note, we did find that many of the submitted saliva specimens from NR were excessively mucoid, clearly containing material that had been coughed into the specimen and had to be digested, similar to the process used for sputum. Following this experience, we issued strict instructions not to cough or do anything that would result in excess mucus in the sample, but to dribble or gently spit into the collection tube. In some other publications the term “saliva” has been used loosely and on reading the specimen description carefully, has clearly included the deliberate collection of mucus. It is unclear from descriptions in most of the published papers where saliva has been studied, however, if they were truly saliva specimens or if they were a combination of saliva and “sputum” and what the relative sensitivity of the two specimen types is for detecting SARS-CoV-2. Saliva does present an attractive alternative to both NPS and NS specimens because the only collection consumable needed is a tube. Notably this sampling strategy has limitations: it may not be suitable for children, mentally challenged patients, or severely ill patients. The collection of saliva is clearly not suitable for intubated patients and the use of alternative sample types should be continued for those situations.

Saliva contains many antibodies and enzymes that could ultimately affect the stability of virus particles or viral RNA, which could impede detection. In our sample comparison from the NR cohort, significantly lower Ct values were observed in saliva samples than those in NS or NPS. As a result, concerns were raised regarding viral stability in this sample type and a series of experiments were undertaken to assess this and the possibility of improving stability with solutions from commercial saliva collection kits. Overall, we found a surprising virus stability in saliva alone for up to 7 days, even at room temperature, whereas the addition of stabilization solutions had adverse effects. It must be noted that these solutions are intended for stabilizing DNA not RNA, and subsequent versions from the manufacturers that are currently being prepared specifically for RNA stabilization may have very different results. It must also be noted that virus stability was only measured with regard to molecular detection. For investigators interested in studies on culturable infectious particles, the stability of virus infectivity may be different.

In symptomatic COVID-19 patients, SARS-CoV-2 viral RNA can be detected one to two days prior to symptom onset and several days post-onset. Thereafter, viral loads decrease substantially, however, there are numerous reports of patients remaining RNA positive for prolonged periods of time and some individuals having intermittent bouts of repeated positivity. Whether these are re-infections, reactivations or simply low levels of residual virus at the limits of detection of the assay, is unclear. Moreover, we have much to learn about the behavior of viral loads in the large numbers of asymptomatic cases now being detected during mass screening of personnel and pre-procedure patients. At this time there is almost no data available on the relative sensitivities of various specimen types in asymptomatic individuals and they may be quite different from the symptomatic populations tested and reported on so far. Thus, the findings of our study, that the combination of NS and saliva provide similar sensitivity to NPS while alleviating many of its limitations, should be carefully considered based on the setting and the study population being tested.

## Data Availability

The authors declare that data that support the findings of this study are available from the corresponding author upon reasonable request.

## Acknowledgements

The authors thank 23andMe^®^ and Ancestry^®^ for their generous provision of the saliva collection kits used in this work, participation in discussions regarding the experiments and their scientific openness regarding the findings. We also thank the staff of the Virology Laboratory at the Wadsworth Center for processing and testing the additional 1500 specimens for the initial phase of this study at a time when testing services were already stretched to capacity. The authors thank Scott Wies, Peter Inglis, Akshay Vig, Kevin Flatley, and Druv Patel for assistance with specimen collection at the AMC site, as well as the nurses and aids who staffed and collected specimens in the AMC tent. We also thank Alan Antenucci for coordinating retrieval of samples from the NR collection site and Bridget Anderson, Rebecca Goldberg, Dominick Chiumento, Richard Cook and Ian Kerr who coordinated the NR sample collection operation.

## Conflict of Interest Disclosures

SBG, GVS, DE, TY, DAC, ACW, AKC, and MJW declare no conflict of interest. KS has served as a paid consultant for T2 Biosystems and Roche Diagnostics. KSG receives research support from ThermoFisher for the evaluation of new assays for the diagnosis and characterization of viruses, including SARS-CoV-2. She also has a royalty generating collaborative agreement with Zeptometrix.

## Funding/Support

This project was performed with internal funding from the NYSDOH and AMC.

## Notes

### Funding Statement

This project was performed with internal funding from the New York State Department of Health and Albany Medical Center.

## REFERENCES

1. Patel R, Babady E, Theel ES, Storch GA, Pinsky BA, St George K, Smith TC, Bertuzzi S. 2020. Report from the American Society for Microbiology COVID-19 International Summit, 23 March 2020: Value of Diagnostic Testing for SARS-CoV-2/COVID-19. mBio 11:e00722–20.

2. Newton PN, Bond KC, signatories from c. 2020. COVID-19 and risks to the supply and quality of tests, drugs, and vaccines. Lancet Glob Health 8:e754–e755.

3. Pere H, Podglajen I, Wack M, Flamarion E, Mirault T, Goudot G, Hauw-Berlemont C, Le L, Caudron E, Carrabin S, Rodary J, Ribeyre T, Belec L, Veyer D. 2020. Nasal Swab Sampling for SARS-CoV-2: a Convenient Alternative in Times of Nasopharyngeal Swab Shortage. J Clin Microbiol 58.

4. Rowan NJ, Laffey JG. 2020. Challenges and solutions for addressing critical shortage of supply chain for personal and protective equipment (PPE) arising from Coronavirus disease (COVID19) pandemic - Case study from the Republic of Ireland. Sci Total Environ 725:138532.

5. Vermeiren C, Marchand-Senecal X, Sheldrake E, Bulir D, Smieja M, Chong S, Forbes JD, Katz K. 2020. Comparison of Copan ESwab and FLOQSwab for COVID-19 Diagnosis: Working around a Supply Shortage. J Clin Microbiol 58.

6. Wang X, Wu W, Song P, He J. 2020. An international comparison analysis of reserve and supply system for emergency medical supplies between China, the United States, Australia, and Canada. Biosci Trends doi:10.5582/bst.2020.03093.

7. Boskoski I, Gallo C, Wallace MB, Costamagna G. 2020. COVID-19 pandemic and personal protective equipment shortage: protective efficacy comparing masks and scientific methods for respirator reuse. Gastrointest Endosc doi:10.1016/j.gie.2020.04.048.

8. Perkins DJ, Villescas S, Wu TH, Muller T, Bradfute S, Hurwitz I, Cheng Q, Wilcox H, Weiss M, Bartlett C, Langsjoen J, Seidenberg P. 2020. COVID-19 global pandemic planning: Decontamination and reuse processes for N95 respirators. Exp Biol Med (Maywood) doi:10.1177/1535370220925768:1535370220925768.

9. Rimmer A. 2020. Covid-19: Experts question guidance to reuse PPE. BMJ 369:m1577.

10. Radbel J, Jagpal S, Roy J, Brooks A, Tischfield J, Sheldon M, Bixby C, Witt D, Gennaro ML, Horton DB, Barrett ES, Carson JL, Panettieri RA, Jr., Blaser MJ. 2020. Detection of Severe Acute Respiratory Syndrome Coronavirus 2 (SARS-CoV-2) Is Comparable in Clinical Samples Preserved in Saline or Viral Transport Medium. J Mol Diagn doi:10.1016/j.jmoldx.2020.04.209.

11. Rodino KG, Espy MJ, Buckwalter SP, Walchak RC, Germer JJ, Fernholz E, Boerger A, Schuetz AN, Yao JD, Binnicker MJ. 2020. Evaluation of Saline, Phosphate-Buffered Saline, and Minimum Essential Medium as Potential Alternatives to Viral Transport Media for SARS-CoV-2 Testing. J Clin Microbiol 58.

12. Rogers AA, Baumann RE, Borillo GA, Kagan RM, Batterman HJ, Galdzicka M, Marlowe EM. 2020. Evaluation of Transport Media and Specimen Transport Conditions for the Detection of SARS-CoV-2 Using Real Time Reverse Transcription PCR. J Clin Microbiol doi:10.1128/JCM.00708-20.

13. Alizargar J, Etemadi Sh M, Aghamohammadi M, Hatefi S. 2020. Saliva samples as an alternative for novel coronavirus (COVID-19) diagnosis. J Formos Med Assoc doi:10.1016/j.jfma.2020.04.030.

14. Azzi L, Carcano G, Gianfagna F, Grossi P, Gasperina DD, Genoni A, Fasano M, Sessa F, Tettamanti L, Carinci F, Maurino V, Rossi A, Tagliabue A, Baj A. 2020. Saliva is a reliable tool to detect SARS-CoV-2. J Infect doi:10.1016/j.jinf.2020.04.005.

15. LeBlanc JJ, Heinstein C, MacDonald J, Pettipas J, Hatchette TF, Patriquin G. 2020. A combined oropharyngeal/nares swab is a suitable alternative to nasopharyngeal swabs for the detection of SARS-CoV-2. J Clin Virol doi:10.1016/j.jcv.2020.104442:104442.

16. Perchetti GA, Nalla AK, Huang ML, Zhu H, Wei Y, Stensland L, Loprieno MA, Jerome KR, Greninger AL. 2020. Validation of SARS-CoV-2 detection across multiple specimen types. J Clin Virol 128:104438.

17. Sullivan PS, Sailey C, Guest JL, Guarner J, Kelley C, Siegler AJ, Valentine-Graves M, Gravens L, Del Rio C, Sanchez TH. 2020. Detection of SARS-CoV-2 RNA and Antibodies in Diverse Samples: Protocol to Validate the Sufficiency of Provider-Observed, Home-Collected Blood, Saliva, and Oropharyngeal Samples. JMIR Public Health Surveill 6:e19054.

18. Wang WK, Chen SY, Liu IJ, Chen YC, Chen HL, Yang CF, Chen PJ, Yeh SH, Kao CL, Huang LM, Hsueh PR, Wang JT, Sheng WH, Fang CT, Hung CC, Hsieh SM, Su CP, Chiang WC, Yang JY, Lin JH, Hsieh SC, Hu HP, Chiang YP, Wang JT, Yang PC, Chang SC, Hospital SRGotNTUNTU. 2004. Detection of SARS-associated coronavirus in throat wash and saliva in early diagnosis. Emerg Infect Dis 10:1213–9.

19. Xie C, Jiang L, Huang G, Pu H, Gong B, Lin H, Ma S, Chen X, Long B, Si G, Yu H, Jiang L, Yang X, Shi Y, Yang Z. 2020. Comparison of different samples for 2019 novel coronavirus detection by nucleic acid amplification tests. Int J Infect Dis 93:264–267.

20. Seaman CP, Tran LTT, Cowling BJ, Sullivan SG. 2019. Self-collected compared with professional-collected swabbing in the diagnosis of influenza in symptomatic individuals: A meta-analysis and assessment of validity. J Clin Virol 118:28–35.

21. Wenham C, Gray ER, Keane CE, Donati M, Paolotti D, Pebody R, Fragaszy E, McKendry RA, Edmunds WJ. 2018. Self-Swabbing for Virological Confirmation of Influenza-Like Illness Among an Internet-Based Cohort in the UK During the 2014-2015 Flu Season: Pilot Study. J Med Internet Res 20:e71.

22. To KK, Lu L, Yip CC, Poon RW, Fung AM, Cheng A, Lui DH, Ho DT, Hung IF, Chan KH, Yuen KY. 2017. Additional molecular testing of saliva specimens improves the detection of respiratory viruses. Emerg Microbes Infect 6:e49.

23. Pasomsub E, Watcharananan SP, Boonyawat K, Janchompoo P, Wongtabtim G, Suksuwan W, Sungkanuparph S, Phuphuakrat A. 2020. Saliva sample as a non-invasive specimen for the diagnosis of coronavirus disease-2019 (COVID-19): a cross-sectional study. Clin Microbiol Infect doi:10.1016/j.cmi.2020.05.001.

24. To KK, Tsang OT, Chik-Yan Yip C, Chan KH, Wu TC, Chan JMC, Leung WS, Chik TS, Choi CY, Kandamby DH, Lung DC, Tam AR, Poon RW, Fung AY, Hung IF, Cheng VC, Chan JF, Yuen KY. 2020. Consistent detection of 2019 novel coronavirus in saliva. Clin Infect Dis doi:10.1093/cid/ciaa149.

25. Centers for Disease Control and Prevention RVB, Division of Viral Diseases. CDC 2019-nCoV Real-Time RT-PCR Diagnostic Panel, Acceptable Alternative Primer and Probe Sets.

26. Centers for Disease Control and Prevention RVB, Division of Viral Diseases. 2020. Research Use Only 2019-Novel Coronavirus (2019-nCoV) Real-time RT-PCR Primers and Probes.

27. Centers for Disease Control and Prevention RVB, Division of Viral Diseases. 2020. CDC 2019-Novel Coronavirus (2019-nCoV) Real-Time RT-PCR Diagnostic Panel, Instructions for Use.

28. Robin X, Turck N, Hainard A, Tiberti N, Lisacek F, Sanchez JC, Muller M. 2011. pROC: an open-source package for R and S+ to analyze and compare ROC curves. BMC Bioinformatics 12:77.

